# A Systematic Review of Spatial Epidemiological Modeling Approaches Applied During the COVID-19 Pandemic

**DOI:** 10.1101/2025.09.24.25336493

**Authors:** Kayode Oshinubi, Ye Chen, Eck Doerry, Esma S. Gel, Crystal Hepp, Tim Lant, Sanjay Mehrotra, Samantha Sabo, Joseph Mihaljevic

**Affiliations:** School of Informatics, Computing, and Cyber Systems, Northern Arizona University, Flagstaff, Arizona, United States; Department of Mathematics and Statistics, Northern Arizona University, Flagstaff, Arizona, United States; Department of Supply Chain Management and Analytics, University of Nebraska-Lincoln, Lincoln, Nebraska, United States; Pathogen and Microbiome Division, Translational Genomics Research Institute, Flagstaff, Arizona, United States; Office of the Vice President for Research, Knowledge Enterprise, Arizona State University, Tempe, Arizona, United States; Department of Industrial Engineering and Management Sciences, Northwestern University, Evanston, Illinois, United States; Center for Engineering and Health, Northwestern University, Feinberg School of Medicine, Chicago, Illinois, United States; Center for Health Equity Research, College of Health and Human Services, Northern Arizona University, Flagstaff, Arizona, United States

**Keywords:** COVID-19, spatial model, movement model, geographical model, meta-population, ODE, PDE, agent-based model

## Abstract

**Background:** A wide range of epidemiological modeling approaches have been applied to the SARS-CoV-2 pandemic, which presents an opportunity to assess common approaches applied to specific research questions. Spatial models interrogate how heterogeneities and host movement dynamics influence local and regional patterns of disease, issues that were of great interest for understanding and controlling SARS-CoV-2.

**Objective:** Here we present a systematic review of spatial epidemiological modeling approaches of SARS-CoV-2. We describe common themes and highlight unique strategies, providing a foundation for researchers to devise spatial models most appropriate for future pathogens and epidemics. Our review also categorizes the research questions that were addressed with spatial models, highlights parameter estimation techniques, and describes the cyber infrastructure used for model development.

**Methods:** We conducted a systematic review using Web of Science and a standardized set of keywords, followed by thorough examination of abstracts and full texts to determine which studies met our inclusion criteria. To guide our description and comparisons of models, we developed a Geography, Population, Movement (GPM) framework that conceptualizes the interactions between three distinct subcomponents of any spatial model. The geographic model represents the physical arena in which the model is implemented, the intra-population model describes the transmission and disease processes that occur within distinct spatial units of the geography, and the movement model describes the algorithms that dictate how hosts move among spatial units within the geography.

**Results:** The search identified a total of 193 articles, of which 109 were included in our review. The most abundant intra-population modeling methods were agent-based (47.7%) and compartmental modeling (29.4%) approaches. Movement models ranged in complexity, with the most complex models implementing commuter movement among many points of interest in the geographic arena, which were sometimes parameterized by fine-scale mobility data. Geographic models ranged from describing microcosms, such as single classrooms, all the way up to multi-country models. Of the 63.3% of models studies that specified the programming language used, we detected ten different languages, with Matlab and Python being the most frequent, although only 30.6% of studies provided open-access code for their models. We also described eight specialized software systems that were used to construct agent-based or compartment models of COVID-19.

**Conclusions:** Our review identified and characterized a variety of spatial modeling strategies and software that were usefully employed to address many relevant epidemiological questions for COVID-19. Future research is needed to quantitatively assess which modeling approaches are most appropriate in specific situations, to answer specific questions, or to apply to certain disease systems. Moreover, future cyber-infrastructure could help to modularize and standardize modeling approaches, which would increase transparency and reproducibility, and which would facilitate a detailed examination of which model attributes relate to model performance in a variety of contexts.

## 1 Introduction

Since the beginning of the SARS-CoV-2 outbreak in late 2019, there have been almost seven million COVID-19-related deaths worldwide, with over 750 million confirmed infections, leading to an astronomical burden on the global public health system [1]. The complexities of a novel emerging pathogen in the context of a globalized society fueled the immediate need for epidemiological studies to evaluate whether non-pharmaceutical interventions could contain transmission, to grapple with the evolution of novel variants, to strategize the heterogeneous deployment of vaccines - Oxford–AstraZeneca, Pfizer–BioNTech, Moderna, Janssen (Johnson & Johnson) and Novavax, and approximately thirty other vaccines that have been authorized in at least one country [2] - and to understand and mitigate growing issues of health inequity [3, 4]. In response, a plethora of mathematical modeling approaches has been deployed to investigate the dynamics of viral transmission and morbidity, evaluate comprehensive intervention strategies, and provide scenario projections and forecasts, leading to thousands of published scientific articles worldwide.

Recent studies have reviewed the vast modeling literature to glean lessons from the COVID-19 pandemic and to suggest future directions. For instance, Kong et al. [5] analyzed the compartmental model structures (e.g., sets of ordinary differential equations) used in COVID-19 dynamic models, including Susceptible-Exposed-Infectious-Recovered (SEIR)-type models that were expanded to consider hospitalizations and the effects of interventions (e.g., quarantined and vaccinated individuals). Other studies[6, 7] surveyed spatiotemporal statistical models to explain how human mobility intervention policies can mitigate a spike in disease transmission. A review by Zino and Cao [8] focused primarily on the development and use of network-based modeling approaches for COVID-19, including dynamic networks, which can take into account time-varying human interactions. For instance, Chin et al. [9] analyzed inter-community networks spatially based on monthly train and bus transportation data, which will be useful for any emerging and re-emerging pathogens. The study revealed the influence of human movements on the spread of diseases. In another related study by Kan et al. [10], the measurement of COVID-19 risk and mobility-based exposure using spatial networks is presented. The study reveals how disparities in human movement are different among sub-populations. These spatial disparities are also presented in [11–13]. Moreover, Xi et al. [14] reviewed papers that used methods from the study of geography to understand COVID-19 transmission and disease impacts, although these methods were largely statistical. Inspired by these insights, our study seeks to specifically review the spatial epidemiological modeling techniques developed and deployed during the COVID-19 pandemic. Spatial epidemiological models are those that implicitly or explicitly represent different host populations within a spatial context, allowing for the movement of hosts to influence local and regional epidemiological patterns [15–17].

Spatial models offer the opportunity to explore important aspects of epidemiological theory, such as the effects of spatial heterogeneities or host movement across disparate populations. For example, Pei et al. [18] developed a spatial model of influenza at the state-level in the USA, parameterizing realistic human movement between states from census data, and this movement influenced when the pathogen arrived in different states. In particular, the authors showed that realistic representation of mobility dynamics within a spatial model better explained the timing of influenza outbreaks across states compared to non-spatial, state-specific models. In a related study, a theoretical article of influenza transmission in Great Britain showed how representing more realistic, daily commuting movement in a model can slow the spatial diffusion of influenza compared to random movements [19]. Such findings underscore the significance of spatial models, highlighting that host movement dynamics can qualitatively and quantitatively impact model-based inferences and forecasts.

Spatial epidemiological models can be highly complex and realistic representations of disease systems. Although capturing this complexity in epidemiological models can present challenges in developing the required computational architecture and analyzing the models [20, 21], there is good evidence that such more realistic models could also improve forecasting accuracy. For instance, the global epidemic and mobility (GLEaM) model [22] simulates the spread of flu epidemics at the global scale and integrates complex data on human mobility from socio-demographic data sets at small spatial scales and from international air transport data sets. Such GLEaM-type models have been notably successful in providing accurate forecasts for influenza dynamics in the USA [23, 24]. Moreover, there are spatial models that include travel between cities via flights can even include transmission dynamics within the aircraft [25].

Designing efficient and tractable spatial epidemiological models requires that researchers make careful decisions about the nature and granularity of spatial characteristics and movement to represent in spatial models, parsimoniously focusing on those with clear impact on overall disease dynamics at different spatial scales, in order to optimize the mathematical and computational complexity of their models. Our aim in this review is twofold: first, to examine the overarching structures of spatial models applied to SARS-CoV-2 and COVID-19, emphasizing assumptions about transmission and human movement dynamics; and second, to assess the computational methods and software systems utilized. While Wang et al. [26] recently reviewed spatiotemporal models for COVID-19—categorizing them into macro-dynamic (like meta-population models) and micro-dynamic models (such as cellular automata), and identifying different data sources for model construction and parameterization. We classify models based on our own modular framework that characterizes spatial models in terms of three fundamental components: (1) the intra-population modeling strategy, which captures transmission and disease processes within populations, (2) the representation of human movement dynamics, and (3) the geographical contexts in which the models were implemented, i.e., the geographic attributes of the spatial arena in which the model is deployed. We also outline the different theoretical and practical scientific questions that were considered across studies, and, finally, we review software that was either previously available and customized for COVID-19, or that was newly designed and released specifically for COVID-19. Our overall aim is to help researchers understand the design of contemporary spatial epidemiological models and understand which methodological choices might be most appropriate depending on the motivations of their future research.

## 2 Methods

### 2.1 Database source and searches

We conducted a systematic search of the literature on spatial epidemiological modeling employed during the COVID-19 pandemic. The following terms and operators were used for a search of abstracts in Web of Science: (covid^*^ OR sars-cov-2) AND (spatial OR mobility OR meta-population^*^) AND (“agent-based” OR “mathematical model” OR “dynamic^*^ model” OR “differential equation^*^” OR “computational^*^ model”). We limited the publications to those appearing between January 1, 2020 and February 1, 2023, including peer-reviewed journal articles or book chapters. In Fig. 1, we present the flow chart of literature search using the PRISMA review statement [27].

**Figure 1:**
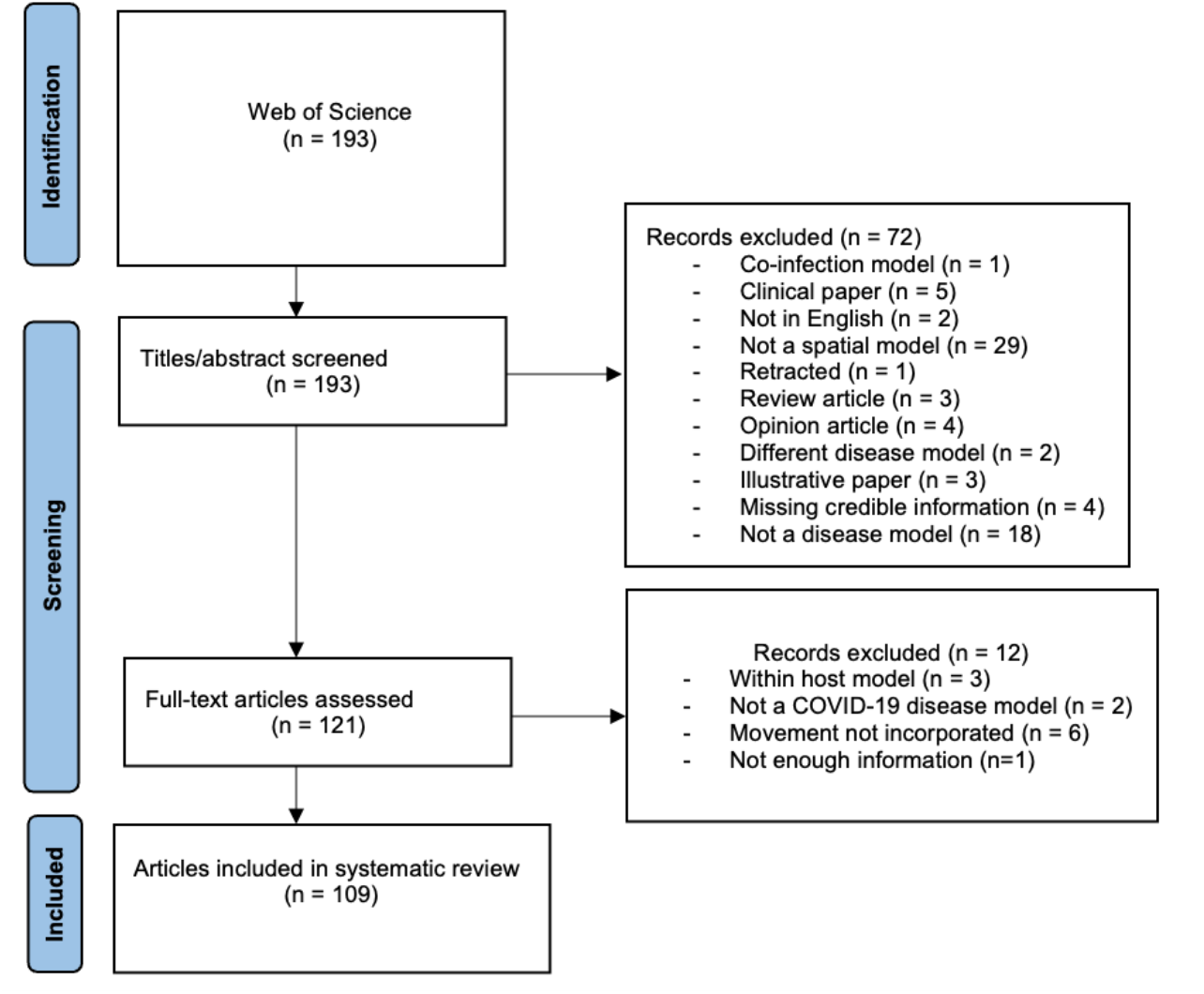
Flow chart of literature search using the PRISMA review statement [27].

### 2.2 Study selection strategy

We included articles in our review based on our definition of spatial, mechanistic models of SARS-CoV-2 transmission and/or COVID-19 progression. Briefly, mechanistic models of disease transmission are those that explicitly represent how susceptible individuals become infected (i.e., the transmission process). A classic example is the standard Susceptible-Infectious-Removed (SIR) compartmental model, which is a set of differential equations that describe the flow of susceptible hosts to an infectious class. This is mechanistically explained by assuming these susceptible hosts contact infectious hosts, and then there is a probability of transmission. More details on mechanistic models are provided in the “Intra-population models (IPM)” section of the Results. Therefore our definition excludes some forms of mechanistic models of population growth, as well as purely statistical models. We briefly cover both of these categories later. Moreover, we were interested in models that represented spatially dynamic systems, which meant they almost invariably included implicit or explicit representations of hosts (i.e., humans) moving between distinct locations (Fig. 2). For instance, we did not consider a standard SIR compartment model to be spatial if it did not include multiple host sub-populations, because such a model would not include spatially explicit movement dynamics (e.g., upper row of Fig. 2) Similarly, we did not consider static network models to be spatial either, because individuals within the network have fixed contacts with other individuals, meaning that they do not not move in space and change their inter-individual connections over time. In contrast, we did consider dynamic network models to be spatial, because the changing contact structure could implicitly represent hosts moving in space and connecting with new hosts.

**Figure 2:**
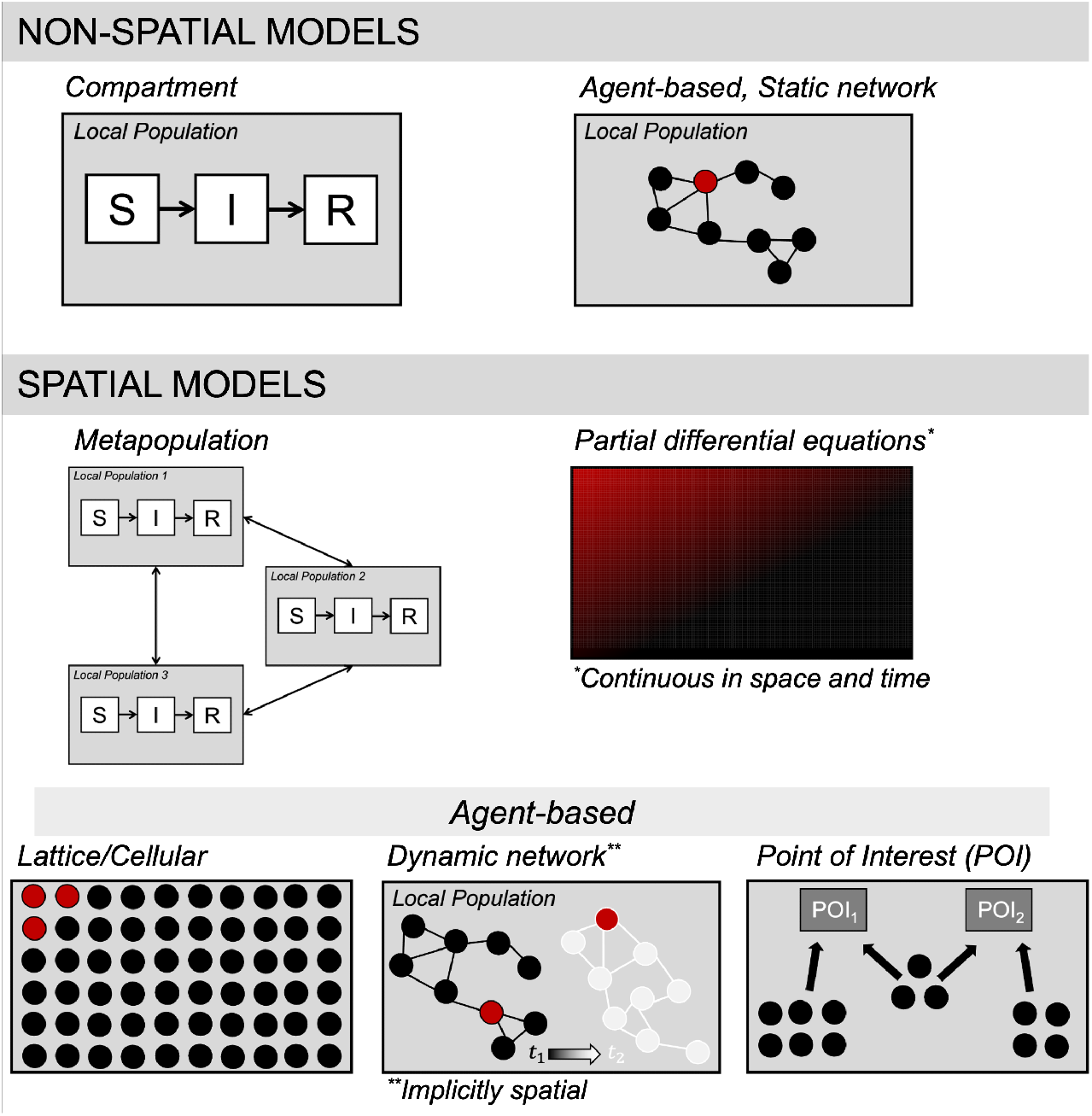
Types of spatial models encountered in our review, highlighting which model types we considered to be “spatial.”

As we describe below, the models we surveyed represent a continuum of models from metapopulation models and continuous-time, continuous-space models, which are defined explicitly by (differential) equations, to agent-based simulation models, in which agents’ dynamics follow certain rules, not necessarily dictated by explicit (differential) equations.

We excluded articles that were not written in English, as well as those in which full-text versions were not available. Also, articles that considered infectious diseases other than COVID-19 were excluded, although we did retain articles that considered multiple diseases (e.g., Cavany et al. [28] studied the effects of COVID-19-related mobility restrictions on dengue transmission).

## 3 Results

The Web of Science search yielded 193 total records. We followed the PRISMA statement to document our selection process[27], an evidence-based, minimum set of items (27-item checklist) developed to characterize and organize systematic reviews (Fig. 1). A total of 109 articles were retained and reviewed. In the Supplement, we provide all of the meta-data that we extracted from the included 101 articles, as well as a list of the articles that were excluded.

### 3.1 Decomposing spatial models into a tractable taxonomy

To structure our results, we will begin by summarizing the model structures that we encountered in the literature (see Fig. 3). One challenge here is that most spatial epidemiological models intertwine multiple aspects of spatial disease dynamics, and these assumptions may be explicit or implicit. In general, there is a description of how the pathogen is transmitted and how disease may progress through the population (e.g., compartments/class, including age classes, infectious classes, hospitalized classes, etc., as well as the rates of transition between classes). For spatial models, there must also be a description of how the host population distributed spatially, along with how local host (sub)-populations are interconnected through host movement. In some cases, two different models may have very similar descriptions of transmission dynamics, but then have very different assumptions about how hosts move through the geographic arena. In another case, for two models, the same compartment model and same movement assumptions may be imposed on two different geographies. And yet, even with their similarities, it can be a challenging task to untangle the explicitly and implicit assumptions any given model and reproduce that model with new code. Clear communication of all aspects of a model is therefore vital for comparing models, for reproducibility and for model validation.

**Figure 3:**
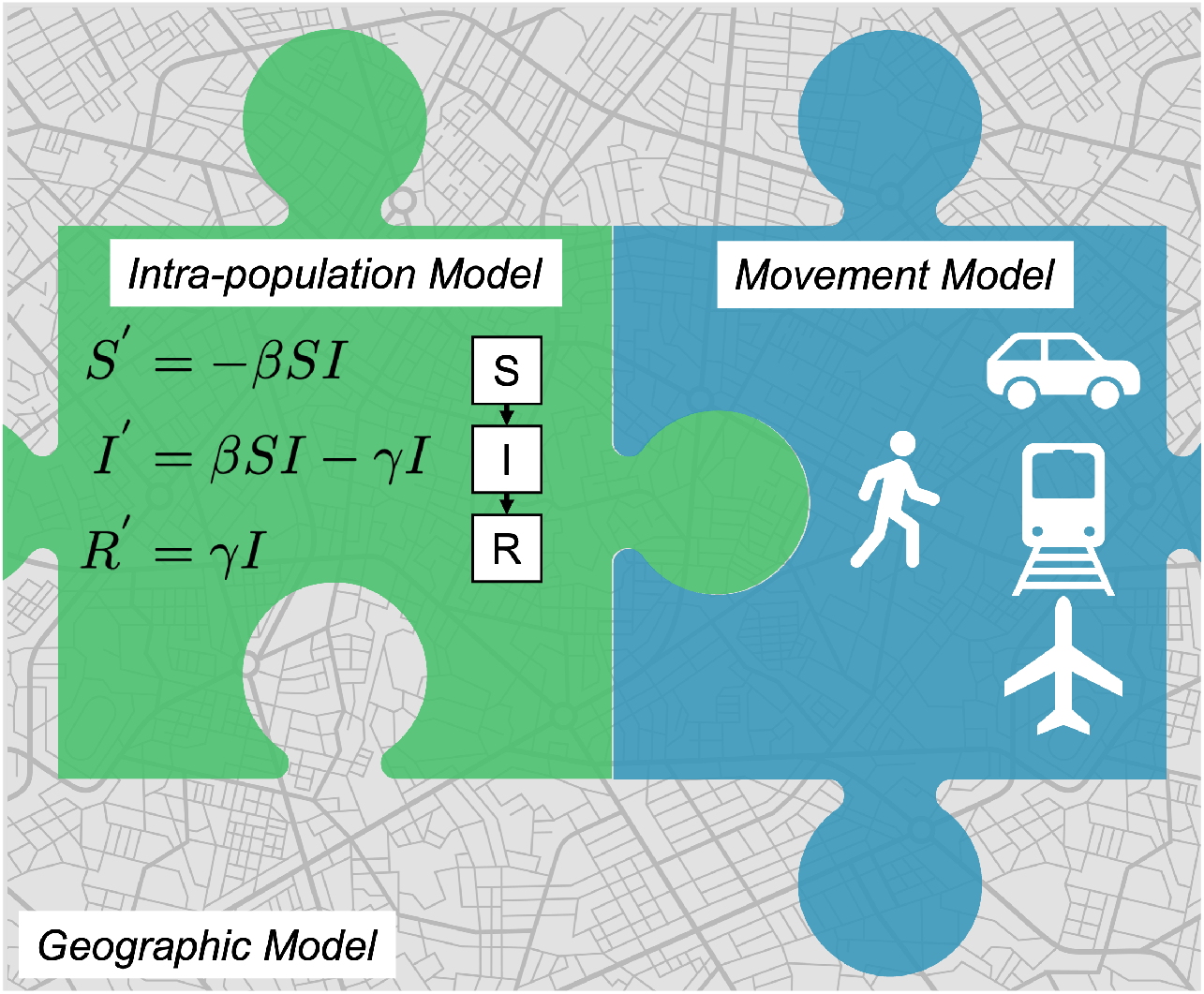
Representation of a spatial epidemiological model using the Geography, Population, Movement (GPM) framework.

As an organizational framework for this review, we develop a descriptive taxonomy called the Geography, Population, Movement (GPM) framework for spatial epidemiological models, which is based on the observation that most spatial epidemiological models can be viewed as the interactions between three distinct elements, or sub-models. An *Intra-population Model (IPM)* describes the transmission and disease dynamics, including model compartments (or, more generally, model processes) and all related parameters, e.g., transmission rates/probabilities). A *Movement Model (MM)* represents the assumptions of how hosts move between spatially distinct sub-populations over time. This could range from a simple gravity model based on centroidal distance between sub-populations, to higher fidelity models that capture travel distance and duration or motivations for movement (e.g. need to access food), to models based on actual observed movements, like mobile device data or airline travel data sets. A *Geographic Model (GEO)* represents all attributes of the geography in which the model is being deployed, including location of population centers, sub-population sizes, local environmental features (e.g., temperature or humidity), demographic features (e.g., social determinants of health), and any other characteristics of local geography required by the IPM and MM. We believe this GPM (Geography, Population, Movement) framework provides a convenient universal framework for characterizing and comparing epidemiological models and modeling approaches. For instance, simpler non-spatial (single population) models can be characterized using GPM as models with a “null” MM, operating in a GEO that described a single host population. In this review, we use GPM to characterize and contrast models developed for COVID-19.

This modular description of models is convenient for comparison, and convenient for computational infrastructure, though we recognize that for some model types these divisions are more conceptual than technical. Below, we propose the question of whether all of the components of a spatial model are perfectly separable. After we describe model structures using the GPM framework and its temporal scales, we discuss the research questions addressed in these studies, as well as the computational infrastructures used to simulate and analyze spatial models. The Supplementary Materials include further discussion of key topics, including how models differed in their time-scales and methods of parameter estimation.

### 3.2 Intra-population models (IPM)

As outlined above, the Intra-Population Model (IPM) describes the processes that influence pathogen transmission and disease progression within a spatially contiguous sub-population. Here the model could be specified by explicit equations, or the model could be governed by a set of rules that are simulated, where the rules may or may not be derived from explicit equations. In Fig. 4, we present the frequency of categories of intra-population models encountered in our review.

**Figure 4:**
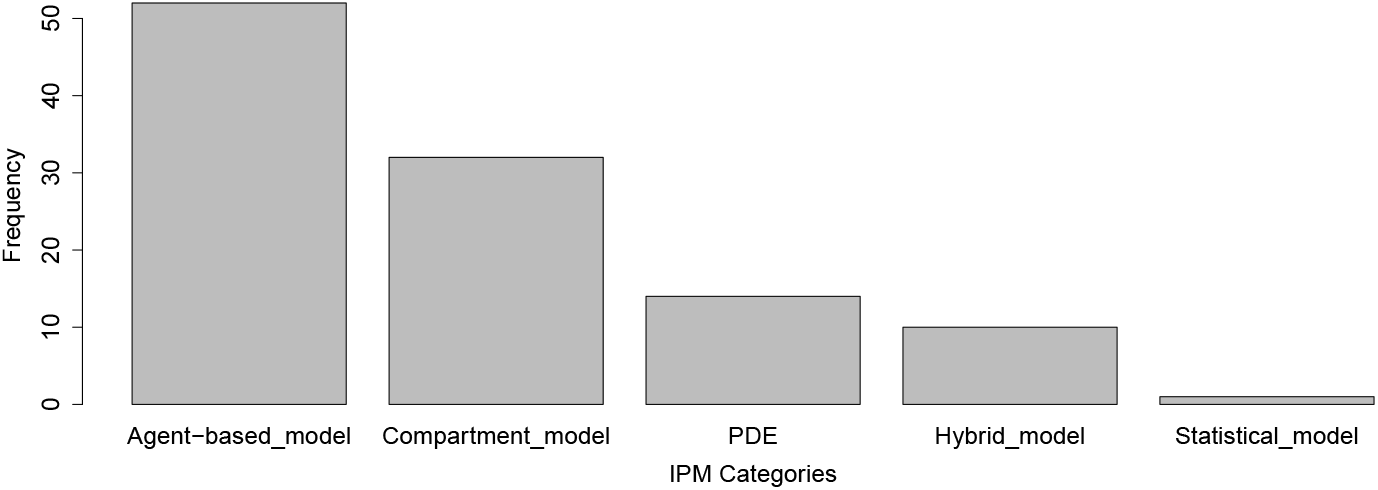
Frequency of categories of intra-population models encountered in our review.

#### 3.2.1 Compartment models with ordinary differential equations (ODEs)

Of the studies analyzed, 29.4% (*n* = 32) employed compartmental models defined by a set of continuous-time equations (e.g., differential equations) to describe disease dynamics within distinct populations. Compartmental models, such as Susceptible-Infected (SI) and its extensions, have been an integral part of understanding epidemiological processes since at least 1760 [29]. Compartmental model structures, in terms of the definitions of state variables, varied significantly across COVID-19 studies, and were recently reviewed [5], so we will avoid a redundant survey of such compartmental structures here. The compartmental models captured key parts of transmission (i.e., that COVID-19 is directly transmitted or close airborne), and disease features (e.g., hospitalization and intensive care).

Briefly, we observed many compartmental model structures that accounted for unique aspects of COVID-19 disease progression. For instance, models such as Susceptible-Asymptomatic-Ill-Hospitalized-Recovered-Deceased (SAIHRD) [30] account for the presence of asymptomatic individuals (see also [31]), as well as delays between infection and hospitalization. Two studies used delay differential equations to better control rates of transition between compartments, such that models could deviate from assumptions of exponentially-distributed rates and better conform to known dynamics of COVID-19, which shows gamma-distributed rates in some cases [32, 33]. Furthermore, it was sometimes assumed that different compartments had different contact rates, such as lower contact rates for hospitalized individuals [34, 35].

Compartment models can also be customized to help evaluate non-pharmaceutical interventions. For instance, models that dealt with testing strategies introduced compartments for detected and undetected individuals [36]. In dealing with the question of quarantining individuals, Topîrceanu [37] developed an SICARQD model (Susceptible-Incubating-Contagious-Aware-Quarantined-Recovered-Deceased), and see [38] for another example. Vaccination was of high interest, and some models added unique compartments for vaccinated individuals. In [39], authors used a compartment of the form Susceptible (S), Exposed (E), Symptomatic (Sy), Vaccinated (V), Recovered (R), and Deceased (D). The study made suggests to decision makers about how current vaccination rates and plans for vaccine expansion could influence the reopening the economy.

#### 3.2.2 Agent-based models

A little above half of the studies (47.7%, *n* = 52) used a type of agent-based modeling to describe disease dynamics. Agent-based models (ABMs) have been used to study the spread of infectious diseases for decades [40]. In ABMs, the disease states of individual agents are tracked through time, and the processes in the model constitute a series of rules that each agent follows (Fig. 2). Unlike compartment models which can be solved using standard numerical methods applied to differential equations, ABMs must use algorithms that dictate the events that befall each individual in the simulation per time step.

While ABMs focus on modeling individuals, the rules followed by agents are often still based on a compartmental disease model. For example, Gomez et al. [41], based their agent-based model on a SEIIIRMD model, standing for Susceptible-Exposed-Asymptomatic Infected (*I*_*A*_)-Seriously Infected (*I*_*S*_)-Critically-Infected (*I*_*C*_)-Immune-Recovered-Deceased. The agent-based approach used by these authors, however, allowed agents’ individual characteristics to alter the overall dynamics, including medical preconditions, age, daily routines (movements between locations), and adherence to social distancing policies. This example highlights that spatial ABMs can be nearly equivalent to spatial compartment models, especially if stochasticity is added to the compartment models. This may beg the question as to whether an ABM is strictly necessary to answer a given question, or if a compartment model is more parsimonious. We address this question in more detail below, however, researchers often use ABMs to add details (e.g., more assumptions or processes) to a model in ways that are more flexible than compartment models. The benefit of ABMs is therefore increased realism that may help explain phenomena that compartment models fail to capture.

The obvious costs are computational complexity and model complexity. Specifically, the complexity of agent-based models arises from the possible set of agent characteristics and the set of rules that agents follow, as well as the ways in which the movement of agents is modeled. See Section 3.2 (Movement Models). The computational complexity can be related to the total number of agents in the simulation, the temporal scale (e.g., how many time steps are advanced), and the number of actions per time step, including transmission processes and movement. This makes ABMs particularly challenging to parameterize, and it can also make ABMs with many agents computationally intractable. Moreover, it can also make interpreting the outcomes of ABMs difficult, especially with respect to reproducibility and identifying which rules or mechanisms are most important [42].

For our purposes of reviewing spatial modeling approaches for SARS-CoV-2, we can differentiate between ABMs that were explicitly or implicitly spatial. Spatially explicit ABMs situate agents into distinct spatial units, or “places”, (e.g., specific buildings or neighborhoods or regions), and agents may move amongst these places. For example, on the low-complexity end of this class of ABMs are cellular automata-type models (including fixed lattices) [43], where agents are placed on grids, and agents can move about the grid. This is considered more simplistic because the grids are usually hypothetical constructs, rather than corresponding to real-life geographies. These models can be useful for generally exploring the effects of individual agent behaviors or characteristics in a spatially heterogeneous landscape.

ABMs can then scale up quickly in complexity, again potentially at the cost of computational complexity and interpretability. Networks (i.e., graphs) can be constructed to represent interactions between agents, where each node is an agent and edges delineate which agents have interactions. Such agent networks can be imposed over realistic geographies [31, 41, 44, 45] to understand the effects of human behavior and mobility. Similarly, ABMs can consider a geographic overlay where spatial units consist of “points of interest” (POI, e.g., grocery stores, parks). As we describe further in Section 3.2 (Movement Models), agents can visit POIs, which creates contacts with other visiting agents, potentially leading to transmission events and spatial diffusion of the pathogen. These latter two types of ABMs are more realistic than, static network ABMs or cellular-automata, but come at the cost of higher computational complexity, and larger dimensionality, requiring more assumptions to parameterize.

Spatially implicit ABMs employ a variety of methods that can effectively represent spatial dynamics without explicitly defining a spatial context. We are considering dynamic network models a form of implicitly spatial ABMs [46, 47], because the connections between agents can change over time, analogous to agents moving in space and contacting new agents in different locations. Dynamic network models for COVID-19 were recently reviewed in [8]. These dynamic network models are of intermediate complexity, because they are clearly more complicated than static network ABMs, but they are not necessarily as complicated as ABMs that incorporate many POI locations.

#### 3.2.3 Spatiotemporally continuous models with partial differential equations (PDEs)

12.8% of studies (*n* = 14) represented spatiotemporally continuous dynamics using partial differential equations (PDEs) [32, 48–59]. PDEs are still compartment models, but spatial PDEs allow for dynamics (e.g., spatial spread of the infection) to proceed in at least two-dimensional space by building these assumptions into the equations (e.g., reaction-diffusion equations [48]). Typically PDEs require numerical methods to simulate the continuous models forward in time, such as finite or meshfree methods, where the spatial dynamics are at least somewhat discretized.

PDEs have advantages such as allowing researchers to represent complex dynamics at small scales with explicit equations, and then integrating these dynamics from small to larger scales. For instance, adaptive mesh refinement and coarsening can resolve population dynamics from local (street, city) to regional (district, state) scales, providing an accurate spatio-temporal description of a spreading pathogen [53]. PDE models are often used to understand the spreading wave dynamics of pathogens, such as in [54], where the authors developed theory on how human mobility affects wave-like spread of COVID-19. A main limitation of the diffusion–reaction PDE approach, however, is that pathogen spread is not only through diffusion to immediate local locations, because people can also travel long distances in short time periods.

Some studies applied novel PDE models to overcome certain limitations of standard compartment models built with ODEs. For instance, Guglielmi et al. [32] presented a new formulation for epidemic models utilizing delay differential equations with PDEs. Oliver et al. [51] developed a PDE model and solved it with a more efficient meshless numerical method, then employed a model reduction technique to construct a surrogate model, aiming to better represent local transmission in France and produced more robust forecasts.

#### 3.2.4 Purely statistical models and Hybrid models

While our review did not focus on purely statistical spatial models, two such studies that passed our initial inspections are notable. Ensoy-Musoro et al. [60] developed a statistical model that explicitly included the dynamic effects of time, space, and movement. Interestingly, Liu et al. [61] compared a non-spatial mathematical model (compartment ODE) to two spatial statistical models (neural network and geographically weighted regression), based on their ability to make forecasts for COVID–19 pandemic in Chinese cities. We also note that Nazia et al. [62] recently conducted a thorough review of the statistical approaches used to understand the spatial and temporal patterns of COVID-19 outbreaks, including methods used to identify socioeconomic, demographic, and climatic drivers. Moreover, we are aware of two other noteworthy statistical models that develop novel methods to account for time and space. Ma and Yang [63] propose an innovative approach to predict the spread of COVID-19 based on internet search volume information (e.g., internet users searching for COVID-19 symptoms, etc.). Additionally, Gayawan et al. [64] used a zero-inflated statistical model with spatial auto-correlation to understand COVID-19 spread across all countries in Africa for the first two months of the outbreak. The model was also able to account for covariates, such as healthcare capacities.

We also identified several manuscripts that combined mathematical models with statistical modeling features within their intra-population model, which we call hybrid models [65–68]. For instance, Wang et al. [69] informed a causal-based graph neural network model with a compartment model to improve spatial forecasting. Interestingly, Yamada and Shi [70] created a non-spatial mathematical compartment model for Tokyo, but then they used PDEs to describe how infected individuals could diffuse from Tokyo to other prefectures, parameterizing location-specific diffusion rates from mobile phone data. A benefit of hybrid models is that they allow flexibility in specifying some mechanisms of the disease or transmission process explicitly, while allowing other features to be described phenomenologically. In some cases this can make it easier to specify, simulate, and/or parameterize the model. But the costs could be easily interpreting the model and forecast accuracy.

#### 3.2.5 Which IPMs are most appropriate for a given research study?

As the examples highlight, the ways to construct the IPM are diverse. Decisions about which IPM to use - whether it is an ODE compartment model, an agent-based model, a PDE - are likely made based on the biology of the host-pathogen system or the specific questions being addressed, although sometimes a researcher’s preference may be a factor. Practical decisions of which IPM to develop may also be based on the modeling tools at the researcher’s disposal, such as access to previously developed model code that can be rapidly adapted, or open-source software tools that could ease the coding of complicated models. For instance, agent-based models were more common in our review compared to compartment-based metapopulation models, but this trend could have been driven by the easy availability of relatively powerful plug-n-play software tool sets for agent-based modeling (see Section 3.6, *Implementation systems, computational environments, and software*). Agent-based models can of course also help to understand the effects of very fine-scale phenomena, such as the effects of transmission between agents while in a commute (e.g., on the bus, train, or airplane). Thus, agent-based modeling could be particularly attractive due to the ease of existing software, as well as the importance of specific fine-grained questions related to person-to-person transmission dynamics of SARS-CoV-2.

Future studies should weigh their choice of IPM against the motivations of the research, the computational complexity of the IPM modeling approach, as well as which data sets are required to parameterize the model’s assumptions. To our knowledge, there has not been a quantitative comparison of IPM model types to understand which are most appropriate in certain situations. For example, agent-based models can capture more fine-scale dynamics of host behavior, including movement dynamics, but, in the case of COVID-19, it is unclear if or when these fine-scale dynamics were necessary to explain observed phenomena or predict future disease dynamics. It is plausible that in some situations a meta-population compartment model, which is more generalized and more computationally efficient, has sufficiently similar performance as an agent-based model within the same geographical location. If so, then the meta-population model would be more conceptually and computationally parsimonious. Future meta-analyses could begin to develop a cogent “map” of the application terrain of epidemiological modeling to clarify which models and approaches are best for addressing which scientific or public health questions at various spatial scales, levels of accuracy, and computational cost. For instance, do agent-based models have higher forecast accuracy of infection dynamics in neighborhoods, but meta-population models are sufficient if you aggregate many neighborhoods into, say, counties? Such quantitative assessments would help emerging researchers better identify the types of modeling approaches that will best fit their needs in terms of speed, accuracy and cost. We believe that researchers should be conscious of how their model structures relate to their study system and specific hypotheses, and, when feasible, deliberately test which model structures best support their hypotheses.

### 3.3 Movement Models (MM)

Various strategies were employed to represent human movement dynamics in spatial models of SARS-CoV-2. In this section, we describe and categorize features of movement models, and then we use our GPM framework to create a classification scheme for movement models along axes of complexity (Fig. 5). Movement models essentially represent theories on why and how host individuals move between locations, and how movement influences spatial diffusion of the pathogen. Therefore, in the same way that some IPMs represent the key processes that influence transmission or disease dynamics within a contiguous population, the assumptions underpinning host movement dynamics can shape how well a model represents important processes contributing to local and regional pathogen spread between population nodes.

**Figure 5:**
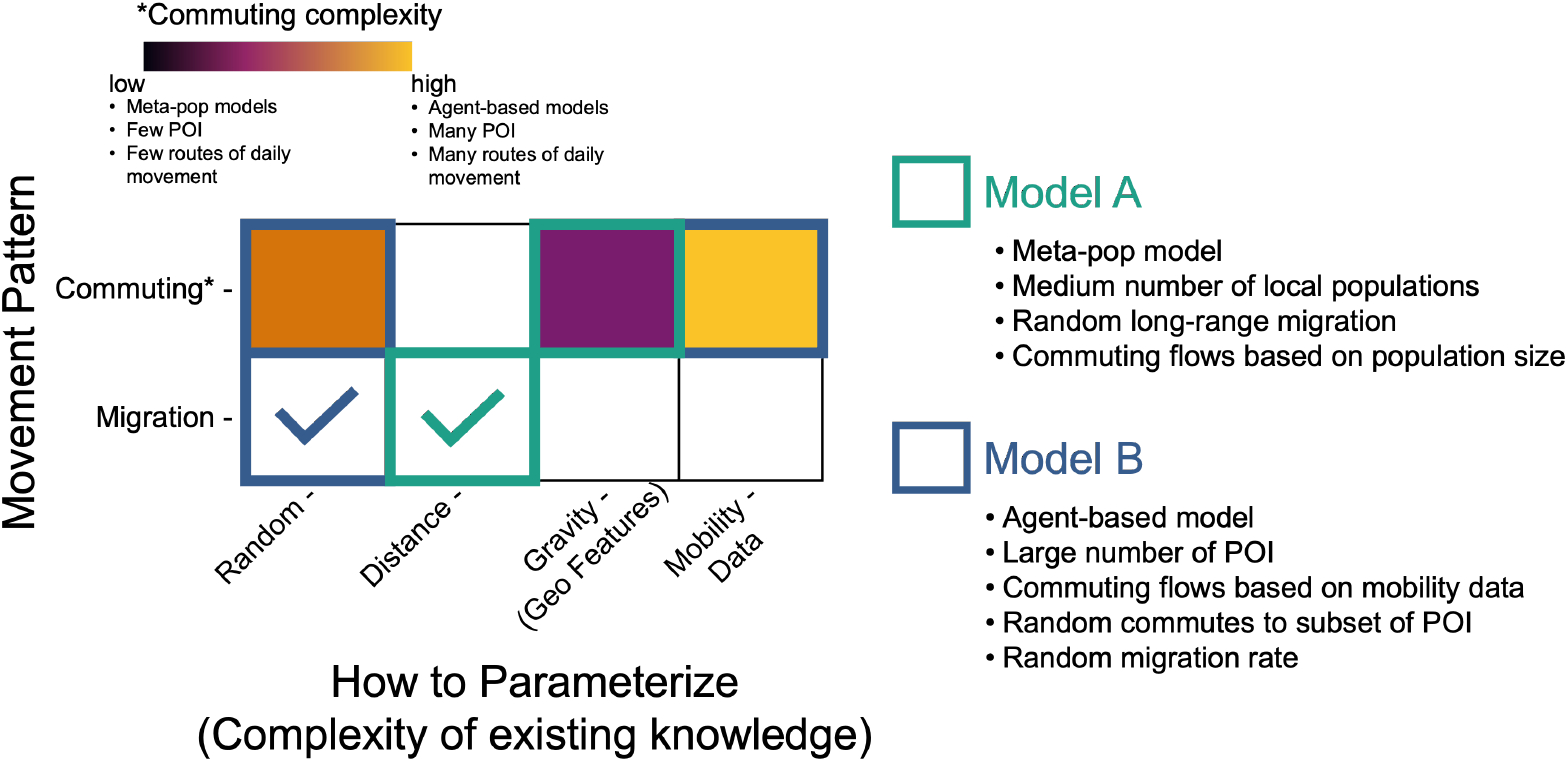
Categorizing movement models along axes of complexity. In this scheme, if commuting dynamics are included, we show that the commuting movement model can range from low-complexity to high-complexity, depending on its assumptions and the types of data used to parameterize the movement. We also distinguish a pattern of migration, where hosts leave their origin, but do not necessarily return. The check-marks show that either a random migration or a distance-based migration rate was included in the example models.

Movement models range in complexity from simple fixed rates of movement, to spatially and temporally fine-scale movement patterns parameterized by high-dimensional data on real commuting flows. The simplest meta-population models can represent movement by incorporating a fixed movement rate between each sub-population within the IPM’s differential equations [33], such that the movement is a continuous-time process. In this case, a very high movement rate causes the infection dynamics of these types of models to become synchronized across the spatial extent, such that the spatial model essentially converges to a non-spatial model. Some computationally complex agent-based models can also employ low-complexity movement models. For instance, ABMs, including cellular automata-type models, can use random-walk movement within bounded areas as a simple way to model movement [37, 71]. In most cases, however, movement dynamics are more realistically linked to features of the geography, or to realistic host behavior. Movement rates between any two sub-populations may be parameterized by features such as population size and points of interest, or movement rates could be impacted by real transportation networks (e.g., roads, train lines, air traffic), or even real mobility data.

#### 3.3.1 Types of movement patterns

##### Distance-based MMs

Distance-based movement models assume that movement is more likely between populations that are closer together in geographical space. This style of movement can be simulated in agent-based models using hard-coded rules or in meta-population models using a method such as a distance-based dispersal kernel that calculates the probability of movement between any two sub-populations as a function of distance [66]. Distance-based movement models can also be parameterized by data in sophisticated ways. For instance, Fazio et al. [72] developed an ABM representing Italy in which movement was parameterized by survey data on distance bands in which individuals routinely move, as well as airline data, demonstrating that movement models can include several categories of complexity.

##### Commuter MMs

We define commuter movement models broadly, because this movement modeling strategy has many unique strategies of implementation. In general, the commuter model assumes individuals (usually a subset of a given local population) move to a new location for some period of time, and then return to their origin location. This commuting movement can represent daily commutes for work, school, or other activities [30, 73, 74]. Some models include routine commuter-style movement in addition to longer-term or permanent movements (i.e., immigration) [71].

To parameterize commuter movement (e.g., how many movers per time per location), sometimes relatively arbitrary assumptions are made about the proportion of the local population that commutes and their duration, while more complex approaches are parameterized with mobility data, such as those generated from mobile devices (e.g., SafeGraph [75]). For example, Keller et al. [36] use mobile device data to parameterize the ratios of people who typically stay at home versus those who regularly commute.

The expansion of mobile device data during COVID-19 led to many models implementing a variation of the commuter movement model, based on *points-of-interest (POI)*, which typically require a fine-grain, agent-based approach. POI movement models can be parameterized with data sources other than mobile devices, however, such as mobility surveys, or these models may be implemented without relying on any specific mobility data (e.g., by making certain assumptions about the frequency of movements). In some cases, agents may be assigned to a household, from which the agents can move to work locations, to shopping centers (e.g., grocery stores), to leisure locations (e.g., theaters and restaurants), or to other agents’ households (e.g., socializing), eventually returning to their own household [39, 49, 76]. Cavany et al. [28] built a city-wide agent-based model and allowed movement to *>* 90, 000 distinct buildings (POIs).

Commuter movement models, including POI, can also restrict which types of movements are allowed during certain times, such as distinguishing between weekday and weekend patterns of movement [77, 78]. In other cases, agents could be divided into school-age, elderly, healthcare workers, etc., and their affinity to certain types of POIs depend on these attributes [79].

Depending on the spatial scales and granularity of the models, some studies distinguish how agents get to their destination (e.g. mode of transit), and may even account for transmission dynamics during this commute. For instance, agent travel via airlines, railways, ships, and cars can be explicitly modeled [39, 70, 79–83]. One model even represented pedestrians explicitly to understand the effects of crowded walkways (e.g., road crossing and sidewalks) on transmission, in addition to vehicular movement [84].

#### 3.3.2 Describing the overall Movement Model

As the above study examples highlight, spatial epidemiological models may use one or more types of movement dynamics, and therefore the overall MM can be described modularly, to account for all types of movement. Figure 5 shows one way we can describe the features of an overall MM, based on the movement pattern and the complexity of existing knowledge that is used to parameterize the model. For example, hypothetical Model A is a low-complexity model in Figure 5; it is a compartment model that has a medium number of sub-populations (i.e., a meta-population model). Model A includes random migration (i.e., hosts can leave to a new place but they don’t necessarily return), and distance between sub-populations influences the number of migrants (e.g., to define long-range migration rates). Model A also has low-complexity commuting movement, specified by a gravity kernel; in this case the kernel equation only includes population size (i.e., hosts are more likely to commute to other populations with large population size, representing population centers).

In contrast, hypothetical Model B has a more complex overall Movement Model. Model B is agent-based, with two types of commuting movement styles. First, mobility data is used to parameterize agent movement between many POIs. Second, there is some random commuting, such that hosts can deviate from their normal commute POI. Model B also includes random migration, in which they move permanently to a new sub-population, but this is not based on any geographic features.

As a practical matter, it was sometimes difficult to discern and clearly identify the movement dynamics and related assumptions used by a modeling study, as movement dynamics were often inter-twined with other model assumptions or within explicit equations that can be difficult to parse. In constructing future spatial models, researchers could use the GPM framework to describe their overall movement model (MM) in a clear, modular fashion. This clear separation and specification of movement dynamics from transmission and disease dynamics (the IPM) could significantly increase the transparency, comparability and reproducibility of modeling experiments.

#### 3.3.3 When are certain MM most appropriate

Modeling the movement of humans for the purposes of an epidemiological model clearly lies along a continuum of complexity. Therefore, it is the challenge of the modeler to determine how much complexity is necessary to address their particular research study. For instance, when is it most necessary to incorporate the large scale data that is needed to implement point-of-interest (POI) movement modeling with an agent-based IPM? These models can be quite complex, so it seems reasonable and helpful to ask when such models are worth the costs of computational and conceptual complexity, as well as the cost of parameterization (i.e., it takes a lot of data to parameterize such models). Unfortunately, there have not been systematic, quantitative research studies to help identify the cases in which certain movement modeling strategies are most appropriate for explaining or forecasting epidemiological phenomena. In our opinion, however, when the research questions are more heavily addressing transmission mechanisms or forecasting disease burden, the movement modeling framework can be less complex. In contrast, if the questions specifically involve where a pathogen might spread in the near future, or which locations may serve as sources or sinks of infection, then the movement modeling strategy would benefit from more detail and data. If the research question is purely a matter of forecasting dynamics, then it seems appropriate to test various modeling strategies and select the ones that most parsimoniously forecast the metrics of interest. Ultimately, we are suggesting that the research question should drive the selection of movement modeling strategy and complexity, and we discuss the types of questions considered by the surveyed studies in Section 3.6 “Research questions addressed”.

### 3.4 Geographical models (GEO)

As outlined earlier, the geographical model (GEO), represents the particular geographical or spatial context in which the epidemiological model was executed. We feel that, the extent and specificity to which spatial models represent the geographic context encompasses a type of model assumption. How a model delineates the local sub-populations of interest can influence the trajectories of the system dynamics; the amount of geographic or demographic detail available in the GEO context dictates what local factors the IPM and MM can consider and incorporate in the dynamics they express. We found many studies that do not have a specific, real-life locations upon which they base their models’ geographies [43, 46, 48, 53–56, 76, 79, 84–95], meaning that their geographic models are essentially implicit, where the GEO contains a few anonymous “places” with limited feature description. In such cases, the focus of the study is to understand general phenomena or generally explore a novel modeling concept, rather than to understand or forecast disease dynamics in specific, real locations. For example, Chondros et al. [96] simulate their model in hypothetical cities, whereas hypothetical small communities were considered by Kevrekidis et al. [48].

For geographic models based on real-world locations, some models are at very small spatial scales (e.g., modeling dynamics in a single building), while others are modeling dynamics that occur among multiple countries, and others include multiple scales in a hierarchy (e.g., counties nested within states). The nomenclature for local populations within the geographic model varied widely, with examples including states, nations or countries, regions or provinces or districts (depending on the nomenclature adopted by the country considered), counties, cities or municipalities or departments, census blocks, continent, electoral wards, and federative units, among others.

The local geographic or demographic information contained in the GEO can be leveraged in various ways by the IPM and MM elements represent that certain processes of infection or movement are dependent on certain local geographic/demographic characteristics. Examples of local geographic characteristics include the size of populations, distances between them, or climate factors like precipitation, temperature, and humidity; demographic factors might include distributions of age gender, or race/ethnicity, or social determinants of health, such as household incomes and employment or healthcare statistics, and so on.

At the smallest scales, many models considered movement between points of interest (POIs), as either attractors situated within geographic sub-populations, or using the POIs themselves as the “places” within which transmission (or even movement) could occur. For example, Gilman et al. [89] looked at how transmission occurred within a medical camp, representing the camp structure with distinct POIs. At larger spatial scales, many studies use established government or administrative boundaries to subdivide locations into distinct populations. This is particularly common when real epidemiological data is used to fit or validate the model, as these data are often aggregated based on such boundaries. For instance, [33] modeled states in India, while 198 urban wards of Bengaluru in India were modeled in [97].

Many studies explicitly included multiple spatial scales within their geographical model [30, 36, 38, 61, 69, 74, 98–111]. For instance, Mhlanga and Mupedza [106] considered movement of individuals between Zimbabwe and South Africa, as well as within Zimbabwean communities. In some cases, custom methods were developed to subdivide real locations for the purposes of representing a geographic model. Two studies divided urban areas (e.g., cities) into square grids, at the 500m scale [78] or the 1 km scale [82].

### 3.5 Are the GEO, IPM, and MM perfectly separable

The choices in designing a spatial model’s IPM, MM, and GEO are of course intricately linked to the motivations and questions being addressed by a particular study (see Section 3.5, *Research questions addressed*). Moreover, the power of an IPM or MM to explain or predict real epidemiological patterns can be fundamentally limited by what information is available in the GEO and can be drawn upon as part of the model’s assumptions. As we showed above, existing modeling studies explore a wide range of spatial conceptions, varying in scale, detail, and what exactly is represented by the “places”, or nodes, in the spatial model. The decisions about which geographic scale, scope and characteristics to capture in a model essentially encapsulate the hypotheses about which geographic features are important to describe transmission processes and explain epidemiological patterns. A researcher’s choice of the granularity of the GEO may be influenced by computational complexity (i.e., how difficult it is to code the model, how long it takes for a simulation to run, etc.), or by the types of data that are available to parameterize aspects of the IPM or MM that depend on the GEO. For example, a researcher may want to build a model representing hosts moving amongst POIs, but the researcher may be limited on the availability of data to parameterize the movement rates to these POIs, or the researcher may want to question whether all types of POI are even relevant to transmission dynamics (e.g., if hosts move to certain types of POI, maybe they are not at risk of transmission, so those POI can be ignored). This example also highlights that the rates of transmission may not be the same at each node (i.e., at each POI), so those assumptions must be determined and described. The choice of GEO may also depend on the availability of epidemiological data. For instance, if data is only available at the State level, and a goal is to fit the model to these data, it may not be straightforward (or even feasible) to validate the assumptions of the model’s processes that occur at sub-State spatial scales.

These issues beg the question as to whether we can truly separate the GEO, MM, and IPM. For instance, some metapopulation models integrate movement into continuous-time, differential equations, such that the movement model and the intra-population model are not technically separate constructs. Or, an agent-based IPM may be coupled with a MM that uses POI, such that this combination of IPM and MM cannot be linked with a GEO scope that does not contain POI. Or, in the previous example, if the model is coded in a way that is not modular, it may be infeasible to apply the same MM and and IPM to a different GEO scope. Therefore, some spatial models may have IPM, MM and GEO that are too intertwined to reliably separate. And, some combinations of IPM, MM, and GEO may not be cross compatible.

Still, it is generalizable, more transparent, and more reproducible to break down the processes and assumptions within an IPM and MM and consider whether those features can be applied equally across different GEO scopes. For example, one may construct an agent-based IPM with a POI-based MM within a single US county, In this case the POI may be intricately tied to the GEO, making it difficult to separate the MM and GEO. However, if this model were specified (i.e., described and coded) generally and modularly, then in theory this same combination of IPM and MM should be transportable to a different GEO. In other words, the model is coded in a way that specifies processes separately from “place”, making it feasible to transpose the same processes onto a different place, provided that POI information is available.

### 3.6 Research questions addressed

In this section, we summarize how the spatial models included in this review were used to understand different aspects of COVID-19 dynamics. Our point is to emphasize how different modeling methods (and different combinations of IPM, MM, and GEO) may be appropriate for certain research questions. The section also demonstrates that often different modeling approaches can address the same questions, and that future work is needed to quantify which modeling strategies are most appropriate for given research questions.

#### 3.6.1 Intervention strategies

Spatial models were used to simulate or infer the effects of various intervention strategies on the spread of the virus, (e.g., interventions specified by the US Centers for Disease Control (CDC) [112]). Some articles focused on *testing strategies*, such as optimizing testing strategies to more effectively identify virus carriers [87], or assessing analyzing the effects of testing policies in relation to mobility restrictions [97]. Spatial models were also useful for studying *vaccination strategies* [85, 113]. Some articles evaluated the optimal allocation of vaccines across localities [114, 115] *or the effects of heterogeneous distribution of vaccinations [116]. Truszkowska et al. [39] examined the interaction between vaccination policies and mobility restriction*.

*Unsurprisingly, spatial models were used to investigate the effectiveness of mobility restriction or social distancing* as an intervention strategy, or tried to predict when mobility restrictions could be lifted [34, 48, 76, 84, 94, 109, 117–119]. Interestingly, Lv et al. [120] used an agent-based model to understand whether traveling in batches, using staggered shifts of commuting, or traveling in isolation had different effects on rates of spatial diffusion of the virus.

#### 3.6.2 Modeling of spatial microcosms

We identified models that attempted to represent highly realistic spatial scenarios, which we refer to as “microcosms”. We chose this term because the models were very detailed and customized to a specific locality, such that the derived insights may only be relevant to other very similar localities. Microcosm models were invariably agent-based. Several studies investigated transmission dynamics within or among classrooms in a school setting [86, 92, 119, 121]. Other models represented university communities, including dynamics within and among dormitories [120, 122]. In a different study, Cuevas [95] considered movement within hypothetical facilities (e.g., movement within a single building). It is unclear if microcosm models are useful beyond their GEO construct. For instance, if we scaled up a university-specific GEO model, and, using its same IPM and MM models, applied them to a whole city, are those IPM and MM equally useful at this new spatial scale?

#### 3.6.3 Individual behaviors or population age-structure

Spatial models were used to explore the effects of individual (i.e., host-level) characteristics or behaviors, or heterogeneities at the population scale, such as different demographic compositions. For SARS-CoV-2 and COVID-19, choices about which behaviors were important depended on the biology of the pathogen (and the biology of pathogenicity), the population of interest, and the policies of intervention. In terms of human behavior, Lombardo et al. [123] examined the effects of social interactions, such as sociability rates, and Guo et al. [110] investigated super-spreading in association with emotional contagion (i.e., the spread of vigilance or panic). Also, Schneider et al. [124] studied the effect of vaccination and COVID-19 variants as it relates to the different age groups in the population. These make sense, as lock-downs and adherence to intervention policies were of concern during the initial outbreak. Some studies considered how population age-structure impacted dynamics, such as age-structured contact rates [125], or age-dependent behaviors and mobility patterns, such as traveling to school [126, 127]. Age-structure was of concern for various reasons, including the observation that older age classes were more susceptible to severe COVID-19, and because questions of whether children could easily get infected and transmit the pathogen were also concerns early in the outbreak.

#### 3.6.4 Questions of general theory

Spatial models were also used to understand how spatial heterogeneities and mobility dynamics can impact COVID-19 patterns in a more general sense. These models provide insights from both a theoretical perspective and for practical applications. For instance, it has been of theoretical interest how long-term dynamics of the virus would be influenced by realistic mobility [128] or seasonal changes in mobility due to climate [108]. Manout and Ciari [129] explored how the combined effects of different daily activities (including movement) might impact transmission dynamics. Spatial models were also used to predict when the virus would reach certain locations via importation from human movement, or the models evaluated the effects of different mobility rates on spreading dynamics [60, 65]. Spatial models were also used to infer the source location of epidemics [46, 47] or to identify hot spots of transmission [66]. These types of “spatial questions” have been asked in the context of many directly-transmitted pathogens, highlighting how spatial models can be co-opted between pathogen systems to ask similar questions.

### 3.7 Implementation systems, computational environments, and software

We noted the computational infrastructures that were used or developed across the studies. Of the 63.3% of studies (*n* = 69) that specified the programming languages used to simulate their models, the most popular programming language was Matlab (*n* = 20) and Python (*n* = 20), followed by C++ (*n* = 6), GAML, which is the language used for the GAMA platform (see below, *n* = 5), R (*n* = 4), NetLogo (*n* = 5), Java (*n* = 3), C (*n* = 2), Julia (*n* = 2), and C# (*n* = 1). Only 33 of the 109 surveyed studies provided open access to their model code. In Fig. 6, we present the frequency of the code availability.

**Figure 6:**
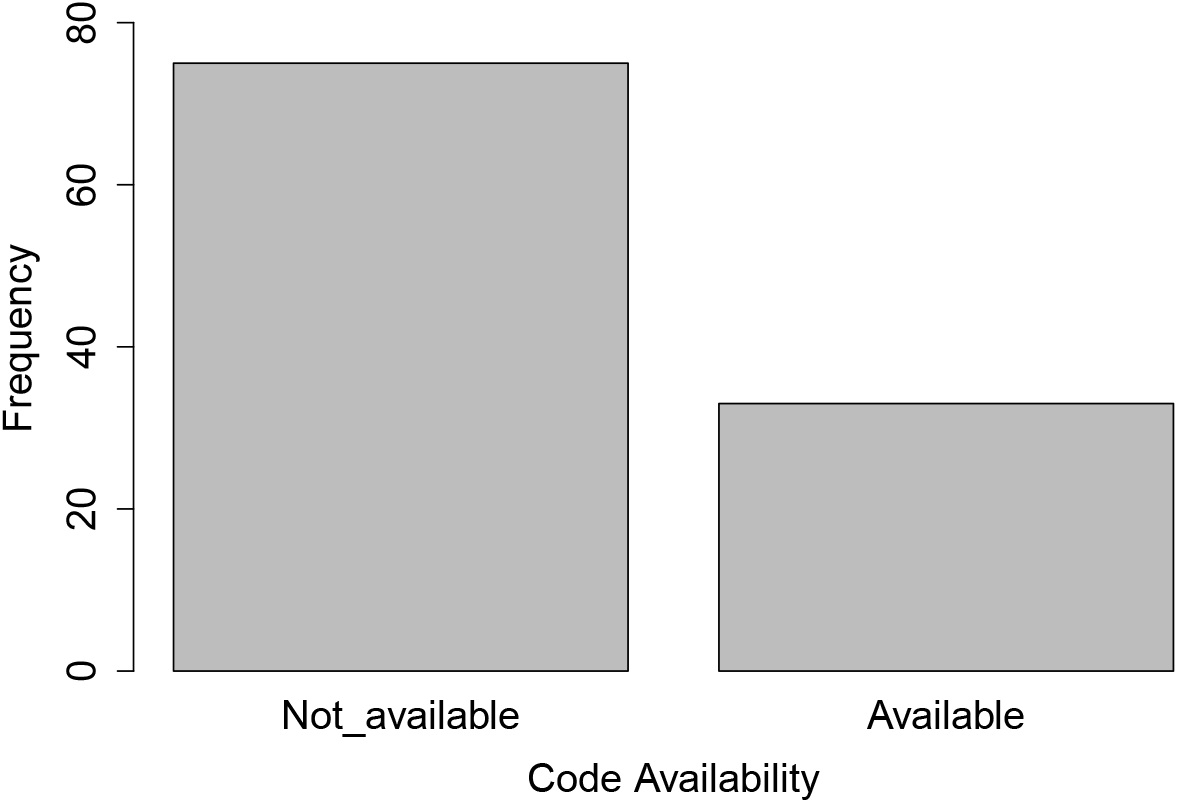
Frequency of literature that made their code publicly available.

Some of the reviewed studies leveraged previously developed modeling software tools to construct models specific to SARS-CoV-2. While we go into more detail in the Supplementary Materials, we note that commonly-used modeling software included GAMA [130], MATSim (Multi-Agent Transport Simulation) [131], and NetLogo [132]. All of these are popular, open-source software for specifying and simulating agent-based modeling studies.

Here, however, we focus on the development of new software to address COVID-19. For instance, PEDSIM [133] is an agent-based modeling platform that was developed specifically for COVID-19, although the original software manuscript did not turn up in our search. The PEDSIM platform is implemented in C++ and simulates transmission dynamics within enclosed spaces, such as a cruise ship or university building, when the floor plan of the building is known. The agents can travel to POIs within the enclosure, and the simulation has rules based on mask-wearing and access to certain areas of the enclosure, to simulate different intervention strategies. Lv et al. [120] used the PEDSIM framework to extend the simulation to an entire university campus and then assess various crowd-management strategies for the control of SARS-CoV-2 transmission.

Covasim is a stochastic agent-based simulator, written in Python, developed specifically for COVID-19 [134]. The platform can provide projections of hospital demand, while also allowing simulations of interventions, such as social distancing, school closures, and vaccination. In our review, Chiba [105] used Covasim to model the entire population of Japan and explore the effects of stay-at-home orders and reducing restaurant business hours.

Pandaesim allows users to simulate stochastic or deterministic versions of an age-structured compartmental model of COVID-19 [30]. The authors implement a customized Gillespie-style algorithm with adaptive tau-leaping, which increases computational efficiency. The system has a desktop version that can be downloaded, where users can customize various aspects of the model simulations, although the geographic model seems constrained to France. Indeed, Pandaesim was used to explore the effects of stay-at-home orders in France, using a meta-population approach [30].

OpenABM-Covid19 is an open source software that allows users to simulate, compare and evaluate different types of interventions, especially vaccination strategies [135]. It is developed using an agent-based epidemic model of COVID-19 disease spread which can simulate all individuals in a given population. OpenABM-Covid19 has R and Python interfaces, but the model is implemented in C for computational (speed) efficiency. In our review, Hinch et al. [73] extended OpenABM-Covid19 to include multiple variants of SARS-CoV-2. The model allows regional variations in the spread of the disease driven by locations spread of variants with non-pharmaceutical interventions (vaccinations).

#### 3.7.1 Limitations of current software systems

As can be seen, almost all of the software systems that were utilized in the reviewed literature dealt with agent-based modeling, several of which were adapted from other research domains to be more relevant to epidemiological studies. There is therefore a notable dearth of standardized and well-tested software architectures for other types of spatial epidemiological modeling, such as meta-population modeling using compartmental ODEs, or implementations of PDEs, at least that we can glean from this review. For these other types of models, this implies that researchers are required to largely develop their own custom code repositories, or rely on lesser-known or validated software tools. And as we show, only a small fraction of custom code repositories are openly shared upon publication. To be fair, custom models within the domain of compartmental ODE models often utilize open-source packages that solve sets of differential equations, but to create spatial models, much more code related to movement dynamics must largely be created from scratch. We are also familiar with some well-known software for network epidemiological models, such as EpiModel [136].

Also lacking are software systems that integrate tools for model simulation and statistical tools for the estimation of model parameters. In other words, while you can build and simulate an agent-based model using some of the reviewed software tools, you will have to supply your own parameter values, and you will not be able to use these software to fit your models to real observational data without developing custom routines. These custom routines can be very difficult to navigate, especially if trying to adapt to new systems, even if the code is open-source. These above issues can clearly hinder the transparency, validation, comparability, and reproducibility of modeling studies.

## 4 Discussion and Conclusion

The aim of our literature survey has been to provide a systematic survey and review of spatial models applied to understand the COVID-19 pandemic. We first developed the Geography-Population-Movement (GPM) framework to provide a uniform framework for characterizing and comparing how various modeling efforts tackled key aspects of the modeling endeavor. This allowed us to modularize the description of spatial models to clarify how each modeling project captured intra-population transmission dynamics, assumptions about host movement dynamics, and the geographical arena considered in the model simulations. We also assessed the implementation systems and software used or built for these spatial models to better understand trends in preferences or conventions. In general, our review highlights the diversity of modeling strategies that have been usefully employed to address a variety of relevant epidemiological questions. Models ranged from simple generalizations of systems that hone in on the effects of specific transmission mechanisms, to highly realistic, agent-based simulations of specific geographical arenas. Specifically, Peng and Liu [137] proposed an agent modeling study that explored spatial local transmission of COVID-19 omicron variant wave in an urban setting across three spatial scales using synthetic mobility structure, demographic, and household characteristics. This modeling study emphasizes how mobility patterns in an urban scaling structure enhance super-spreading of the disease, an approach which is a deviation from the statistical methodology presented by Zou et al. [138] to study COVID-19 variants at an international scale (a study of 100 countries).

An important outcome of our survey study is the need for increased standardization in the conceptual and communicative frameworks used by in modeling studies. In our opinion, having a common basis for articulating key features of a modeling study would help to increase transparency, comparability, and repeatability of modeling efforts, as well as promoting the creation of a growing set of powerful modeling software tools that could substantially streamline the exploration of new modeling ideas. We have attempted to provide a first step in this direction with the descriptive GPM framework, which we used to characterize models and organize this survey. Such a framework would allow modelers to clearly characterize the foci and contributions of their modeling efforts, i.e., theory of transmission dynamics (IPM), host movement dynamics (MM), or leveraging of geographic context (GEO). This would reduce the often intense amounts of analysis (and sometimes guessing) required to untangle and understand the detailed behavior of the many custom-created models we encountered in our review, and thereby accelerate efforts to compare approaches, correctly assess the reasons behind different models producing different patterns, and ultimately better organize our collective efforts towards spatial epidemiological model development. If translated to the implementation level, such a standardized descriptive framework would support the development of powerful modeling software frameworks that allow rapid and focused exploration of a wide range of modeling ideas while reducing the threshold of entry for non-programmers.

Our review strategy purposely focused on mechanistic models that included explicit representation of the transmission process. It is worth nothing that mechanistic population growth models (e.g., sigmoidal or logistic population growth) were often used to understand and predict the growth of the infectious population. While these models do not typically include explicit infection dynamics, and were therefore not formally included in our review of mechanistic disease models, this type of modeling approach is popular and deserves some attention here. For instance, Konstantinov et al. [139] tested several growth models and identified that the generalized fractional-power model and the generalized inverse tangent model fit well to patterns of growing infection case counts in Europe. Similarly, Ahmadi et al. [140] tested three general growth models, the Gompertz, von Bertalanffy, and the cubic polynomial, against COVID-19 data in Iran. Also, El Aferni et al. [141] applied a sigmoidal-Boltzmann model to study the spread of COVID-19 in several countries, allowing them to compare what the authors call the spread rate, which is analogous to the transmission rate. Interestingly, there have been notable recent advances in this growth-based modeling approach. For example, Essadok-Jemai and Al-Rabiah [142] proposed an extension of the logistic growth model that allowed for multiple transitions during the pandemic period (e.g., shifts of increasing or decreasing growth rate of the infectious population), which fit well to data at the global scale.

As this review illustrates, the COVID-19 pandemic in many ways accelerated the development of spatial modeling research, making apparent the critical need to consider the complex movement of hosts in within realistic, heterogenous landscapes. The creativity and range of approaches and ideas this has generated has been impressive, but also emphasizes the need for spatial modelers to move beyond brainstorming to develop a broader shared understanding of the nature and dimensionality of the modeling space that is being explored. Without this and the shared terminology that comes with it, individual studies will remain bright points of interest that are difficult to characterize, compare, replicate, and evaluate with respect to their applicability to different geographic contexts or different disease systems. With this in mind, we attempt to identify future challenges and opportunities in spatial epidemiological modeling.

A key opportunity, but obvious challenge, is the development of a descriptive framework to detail the various assumptions of spatial epidemiological models, from transmission dynamics, to movement dynamcis, to geographical context. Other fields of research have mechanisms to enhance standardization of practice, description, and/or data formatting, such as genomics (e.g., GenBank, GISAID). Recently, the epidemiological modeling community has proposed guidelines for reproducibility in terms of describing a modeling study [143, 144], echoing calls for enhanced transparency across fields that rely on scientific software [145]. Our GPM framework represents an effort in this direction specifically for spatial epidemiological models, which have unique aspects of complexity, but this framework will surely require refinement, extension or even replacement to arrive at a common basis for characterizing a sufficiently inclusive set of spatial modeling approaches.

With a framework capable of clearly characterizing the relative conceptual placement of various modeling approaches within a well-articulated modeling space, one could begin developing at least a heuristic understanding of what modeling approaches are most efficient, accurate, and parsimonious for different types of research questions. Which modeling approaches are most promising for understanding a particular type of pathogen at specific spatial scales? Can we begin to generalize which types of spatial hypotheses are best tested with differing types of spatial models? Exploring these questions quantitatively would require standardized methods for evaluating the performance of spatial models against real-world observational data. At least for model-based forecasting of infectious disease dynamics, the community is being to develop such methods that have emerged from collaborative modeling hubs [146]. Common evaluative approaches and scoring metrics will be key to comparing the performance of differing models and approaches, as well as providing a basis for understanding the cost-benefit ratio offered by each. Similarly, we should develop best practices for spatial model parameterization - or at least more robust and standardized descriptions of how parameterization was accomplished-based on a growing understanding of which optimization or fitting algorithms perform best for which types of models. Relatedly, we can work towards better understanding the spatial scale that is appropriate for estimating location-specific parameters (e.g., determining the scale at which point parameters are no longer “identifiable”) based on available data.

A critical practical challenge for enabling and streamlining exploration of all of these issues is the development of powerful, flexible cyberinfrastructures that allow modelers to rapidly turn novel modeling ideas into implemented runnable models, easily test those models against data in differing geographic places, and compare their model’s performance to others. Such a cyberinfrastructure could potentially be extended from scenario simulation to include forecasting with the future development of tractable statistical approaches and software tools for automating or streamlining parameter estimation for complex spatial models. Numerous software packages are under active development and quite promising, a future scalable spatial modeling environment will play a critical role in the deployment of accurate forecast models that will allow for the robust evaluation of relevant interventions and serve as a guide for effective decision making.

## Data Availability

All data produced in the present work are contained in the manuscript

## Supplementary information

Extensive meta-data on each of the publications we reviewed is added as a supplemental file.

## Declarations

### Funding

Research reported in this publication was supported by the National Institute of Allergy and Infectious Diseases of the National Institutes of Health under award number R01AI168144.

### Competing interests

The authors have no competing interests.

### Ethics approval and consent to participate

Not applicable.

### Data availability

The data used for this research is added as supplementary material.

### Consent for publication

Not applicable.

### Author contribution

Conceptualization, K.O. and J.M..; methodology, K.O. and J.M.; software, K.O.; validation, K.O., Y.C., E.D., E.G., C.H., T.L., S.M., S.S., and J.M.; formal analysis, K.O.; investigation, K.O., Y.C., E.D., E.G., C.H., T.L., S.M., S.S., and J.M.; resources, J.M.; data curation, K.O. and J.M.; writing and original draft preparation, K.O. and J.M.; writing—review and editing, K.O., Y.C., E.D., E.G., C.H., T.L., S.M., S.S., and J.M.; visualization, K.O. and J.M.; supervision, J.M.; project administration, J.M. All authors have read and agreed to the published version of the manuscript.

## Notes

### Competing Interest Statement

The authors have declared no competing interest.

