## Supplemental File 1 for "A Systematic Review of Spatial Epidemiological Modeling Approaches Applied During the COVID-19 Pandemic"

Oshinubi et al.

November 8, 2024

In this Supplementary Materials, we consider two additional topics that are relevant for surveying the spatial modeling studies encountered in our review. First, we address differences in the temporal scales of the models. Second, we address the various methods and motivations for estimating parameters of these often complex spatial models. Third, and finally, we describe implementation systems and software that was commonly used to simulate agent-based models, but note that these software are not new or unique to COVID-19.

#### 1 Temporal scales of the models

Thus far, we have used the GPM framework to characterize and compare the structural aspects of published COVID-19 models, i.e., how they differ in their representation of infection dynamics (IPM), movement dynamics (MM), and representation of the spatial context (GEO). Orthogonal to these structural characteristics, existing models can also be characterized by differences in temporal scale, defined as the time steps by which the models are solved or simulated forward. In general, each time step in a spatial model integrates two processes: the “transmission step”, in which the IPM runs a time step to compute transitions between model compartments (i.e., infection or disease classes) within a spatial sub-population (node), and a “movement step” in which the MM computes host displacements between locales (nodes) represented in the GEO. These two steps alternate (or occur simultaneously) throughout the time period of the model execution, effectively “solving” or “simulating” the system dynamics. In many cases, the temporal scales of IPM and MM are identical, but this is not necessarily the case; several finer grained movement steps (e.g., hourly) could be modeled between each infection step (e.g., the IPM runs daily).

80.7% of studies ($n = 88$) used a daily time scale (i.e., 24-hr), and 13.8% of studies ($n = 15$) used an hourly time scale. The frequency of the temporal scales observed in the literature search is presented in Fig. 1. Hourly time scales were often chosen when movement dynamics varied between day-time and night-time (e.g., commuting individuals in their workplace/school or at home). Only one of the studies implemented a weekly time scale [1], while five studies used a minute time scale [2–6]. The reasons why a researcher may choose a particular time scale is not necessarily straightforward. Some existing software dictate which time scale is used in the coding architecture, so it is outside of the user’s hands. In other cases, rates in the model may have been estimated from data that is collected at certain time scales, so a time scale is chosen to maintain the same units of measure. There could be reasons to suspect that transmission or movement processes occur at specific time scales that influence local or regional diffusion of the pathogen. Or, it could be the case that epidemiological data are reported at certain time frequencies, so it is convenient to match the model time-scale to those frequencies.

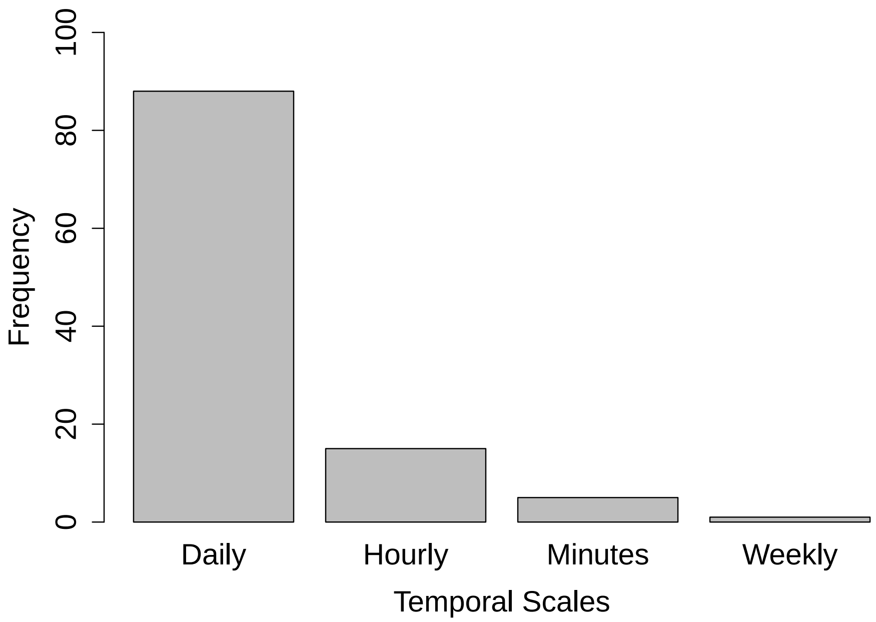


**Figure 1:** Temporal scales observed in the literature search.

#### 2 Parameter Estimation and Forecasting

To accomplish the study motivations discussed above, it is often necessary to parameterize models (i.e., estimate the parameter values from real data). We can divide the goal of parameterization into three domains. First, parameterization could heavily rely on previously published parameter values, such that the model simulations are at least reasonable approximations of dynamics that could be observed. Second, parameters may be fit to recent or historical data to understand if the model can match those data well (i.e., a hindcast), which may indicate that the model has appropriate assumptions to describe the system. Third, parameters may be fit to recent data so that the model is tuned for the prediction of future data under certain assumptions of future conditions (i.e., forecasting). Often, hindcasting is an initial step towards developing a forecast.

In our review, 50.5% of the total studies ($n = 109$) conducted hindcasting ($n = 55$), forecasting ($n = 4$), or both ($n = 8$). These studies used a variety of parameter estimation techniques, which we will cover briefly. Some methods are more quantitative and statistical than others. For instance, many studies ($n = 24$) predominantly relied upon previous knowledge to parameterize the models (e.g., previously published parameter estimates). Another subset of studies used some type of manual tuning ($n = 9$) to estimate some parameters. Manual tuning can be guess-and-check with a qualitative comparison to data, or this could be more quantitative. For instance, while Lombardo et al. [7] chose a limited set of values of a parameter to test against data, they used likelihood metrics to decide which among the parameter set produced model simulations that best matched the data. For seven studies, we could not clearly discern how some of the parameters were estimated.

The other studies employed what we broadly categorize as probability-based or distance-metric-based estimation techniques, which include maximum likelihood, optimization, and Bayesian inference. For instance, four studies used Approximate Bayesian Computation and six studies used formal Markov Chain Monte Carlo (MCMC; full Bayesian) algorithms. One study used particle filtering method to infer parameter estimation at the County scale Calvetti et al. [8]. Eight studies used a version of non-linear least squares estimation, while nine studies used optimization techniques, including two studies [9, 10] that used Optuna [11], a Python-based optimization toolkit.

As a specific example, Wang et al. [12] proposed a novel spatiotemporal learning framework with a causal module that connects the neural network with the compartmental model called causal-based graph neural network, and used the method to forecast daily cases of COVID-19 at global, US state, and US county levels. In [13], Approximate Bayesian Computation was used for model fitting of daily national death cases in Italy, Spain and the UK to estimate four parameters. Yamamoto et al. [14] used a combination of MCMC and regularization to estimate parameters (i.e., hindcast) and then made forecasts for various countries in Europe and for various cities in China. To our knowledge, there have not been systematic studies of which parameter fitting methodologies yielded more precise hindcasts or forecasts. Some studies attempted to improve forecasting by using realistic, fine-scale movement dynamics [15, 16]. Similarly it is unclear if the structure of the IPM, MM, and GEO components of the model have a larger impact on forecast accuracy as compared to which parameter fitting method was used. These are clear questions for future quantitative syntheses of the literature.

### 3 Previously developed implementation systems

Five articles utilized the GAMA [17] software [2, 3, 18–20]. GAMA is an open-source modeling and simulation environment for creating spatially explicit agent-based simulations, and it is not specific to epidemiology. GAMA is written in Java and made of Eclipse plugins, which can be customized to meet specific needs and can be interfaced with other languages (e.g., R or Python). GAMA comes with many options for model visualization, including interactive animations and 3D rendering, and users can overlay agent-based models on custom GIS layers. For example, Gaudou et al. [18] utilized the GAMA platform to create the COMOKIT (COVID-19 MOdeling KIT), which defines agent characteristics and action rates specific to COVID-19, as well as realistic movement actions (e.g., going to work), with the ability to simulate various intervention strategies.

MATSim (Multi-Agent Transport Simulation) [21] is an open-source, Java-based platform that is has been developed for decades, and which simulates the movement of agents along realistic transportation networks within cities. In our review, Manout and Ciari [22] leveraged MATSim as well as the agent-based disease transmission model, EPISim [23], to understand how realistic transportation networks and movement dynamics influence COVID-19 patterns in Montreal, Canada. MATSim was also used to explore the effects of mobility restriction on SARS-CoV-2 in South Africa [24].

NetLogo [25] is an open source software package based on an agent-based programming environment that has been around for two decades and has more recently been adapted to simulate epidemics. While the back-end of NetLogo uses Java, and users can create custom Java-based models, most users rely on NetLogo's custom front-end language or graphical user interfaces that are more user-friendly. In our review, several studies used NetLogo software to simulate their agent-based model [26–29]. For example, Macalinao et al. [28] used NetLogo to build classroom layouts similar to those in the Philippines and explore the effects of altering seating arrangements within classrooms and altering how students rotated classrooms between sessions.
